# Optimal LDCT screening for never-smoking Asian women using integrated polygenic and environmental risk: a microsimulation modelling study

**DOI:** 10.64898/2026.08.18.26360665

**Authors:** Akiko Kowada

## Abstract

**Objective:** To identify optimal initiation ages and screening intervals for low-dose computed tomography (LDCT) screening among never-smoking Asian women using an integrated polygenic risk score (PRS)–environmental tobacco smoke (ETS) risk model, and to evaluate the cost-effectiveness of alternative screening strategies at these optimized ages.

**Design:** Integrated PRS–ETS microsimulation modelling.

**Setting:** Japan.

**Participants:** Never-smoking women stratified into eight risk groups defined by combinations of PRS levels and ETS exposure.

**Interventions:** LDCT screening at intervals of 1 to 10 years, annual chest radiography (CXR), or no screening.

**Main outcome measures:** Costs, quality-adjusted life years (QALYs), incremental cost-effectiveness ratios (ICERs), net monetary benefits, lung adenocarcinoma incidence and mortality, and optimal LDCT initiation ages. Sensitivity analyses used a willingness-to-pay threshold of US$50,000 per QALY gained.

**Results:** Optimal initiation ages ranged from 40 to 55 years across the eight PRS–ETS risk groups, with higher PRS–ETS risk associated with younger optimal initiation ages. Annual LDCT was the most cost-effective strategy across all PRS–ETS risk strata, yielding an ICER of US$40,471 per QALY in the lowest risk stratum and becoming cost-saving in higher risk strata. Over a lifetime, annual LDCT averted 8,534 lung adenocarcinoma deaths compared with annual CXR and 14,940 deaths compared with no screening.

**Conclusions:** Tailoring LDCT initiation age across integrated PRS–ETS risk groups maximizes mortality reduction achievable with cost-effective annual LDCT screening among never-smoking Asian women. These findings highlight an urgent limitation of global lung cancer screening guidelines that rely exclusively on smoking history and provide policy-ready evidence supporting the integration of PRS and ETS into future recommendations for precision LDCT screening for never-smoking populations.

## Introduction

Lung cancer remains the leading cause of cancer death worldwide [1,2]. Tobacco smoking has historically driven most cases, and major screening recommendations were developed around smoking-related risk [2].

Randomized trials of low-dose computed tomography (LDCT) screening for high-risk smokers, including the NLST and NELSON trials, demonstrated mortality reductions and have informed screening policy in several countries [3,4].

By contrast, a growing burden of lung adenocarcinoma among never-smoking women has emerged across East and Southeast Asia [5–7]. In Japan, adenocarcinoma comprises the majority of lung cancers in women (≈82% in national registry data) and is even more common among never smokers (reported up to ≈88% in cohort analyses) [5–7, 8]. These regional increases have occurred despite falling population smoking prevalence, suggesting a shift in epidemiology that is not explained solely by active smoking trends [5, 6, 9].

Existing screening frameworks offer little guidance for never-smoking Asian women. Randomized LDCT trials enrolled predominantly heavy smokers and did not include never smokers, limiting direct evidence for screening effectiveness in this population [3,4]. Observational LDCT studies and risk-prediction efforts among never smokers have provided important insights but have not produced validated, policy-ready risk thresholds for LDCT screening [10–12].

Tumors arising in never smokers often occur in the peripheral lung and can be curable when detected early [10,13]. Nevertheless, many countries in Asia have historically relied on chest radiography (CXR) for opportunistic or population screening; CXR has substantially lower sensitivity than LDCT for peripheral adenocarcinoma and frequently detects disease only at advanced stages, contributing to underdiagnosis and delayed treatment [3, 4. 14].

At the same time, polygenic risk score (PRS) for lung cancer is maturing, and several studies indicate that inherited susceptibility contributes meaningfully to lung adenocarcinoma risk among never smokers [15–18]. Environmental tobacco smoke (ETS) exposure remains common in parts of Asia and is increasingly recognized as a contributor to lung cancer risk in never smokers [19–22]. Integrating PRS-ETS risk information therefore offers an opportunity to develop precision LDCT screening strategies tailored to never-smoking Asian women.

Current guidelines have not incorporated integrated PRS–ETS risk stratification for never smokers, leaving policymakers without clear evidence to update recommendations [14, 23]. As LDCT capacity and interest in screening expand across parts of Asia, defining who to screen, at what age, and at what interval has become a pressing public health priority [14, 24].

To address this evidence gap, we developed an integrated PRS–ETS risk model and applied an individual-level microsimulation to identify optimal LDCT initiation ages and evaluate cost-effective screening strategies for never-smoking Asian women.

## Methods

### 2.1 Model overview

We developed an individual-level microsimulation to estimate optimal initiation ages for LDCT screening among never-smoking Japanese women. Annual LDCT strategies beginning at ages 40, 45, 50, 55, 60 and 65 years were embedded directly into the model to enable consistent comparison across screening intervals and initiation ages. One-way sensitivity analyses varied relative risk continuously from 1.0 to 8.0 to cover the risk range represented by the eight combined PRS–ETS strata and to identify threshold points at which the preferred initiation age for each stratum changed [15, 16, 19, 20]. Optimal initiation age for each stratum was defined as the age that maximized net monetary benefit (NMB) at a willingness-to-pay threshold of US$50,000 per quality-adjusted life year (QALY) gained [25].

After determining stratum-specific initiation ages, we constructed eight independent microsimulation state-transition models representing the four PRS strata crossed with two ETS exposure categories. Each model compared LDCT screening at intervals of 1, 2, 3, 4, 5, and 10 years with (a) annual chest radiography (CXR), representing the current national approach in many settings, and (b) no screening [3, 10,11, 14]. The model structure is generalizable to other East Asian populations and intended to inform national and regional screening policy.

Analyses were conducted from a healthcare payer perspective over a lifetime horizon. A one-year cycle length with half-cycle correction was applied. Costs and QALYs were discounted at 3% per annum. [26]

Primary outcomes were costs, QALYs, incremental cost-effectiveness ratios (ICERs), NMBs, lung adenocarcinoma incidence, lung adenocarcinoma mortality, and optimal LDCT initiation ages. Cumulative lifetime outcomes were estimated using a Markov cohort framework embedded within the individual-level microsimulation. Population-level outcomes were obtained by multiplying per-person differences in costs, QALYs, and lung adenocarcinoma incidence and mortality by the size of each target population; age-specific lifetime effects were applied to the 2024 Japanese female population [27].

### 2.2 Target population

The target population comprised Japanese never-smoking women. Participants were stratified into eight risk groups defined by the cross-classification of four PRS strata and the presence or absence of ETS exposure. PRS strata were defined as quartiles of the validated multi-ancestry/East-Asian PRS distribution (lowest to highest risk). ETS exposure was defined as any reported household, workplace, or frequent public-space passive smoking exposure versus none. Optimal LDCT initiation ages were estimated separately for each risk group. [15, 16, 17, 19, 20, 22, 27]

Never-smoking status was defined as self-reported lifetime consumption of fewer than 100 cigarettes, a standard epidemiologic definition used in international smoking-related research.

### 2.3 Risk stratification

PRS were grouped into quartiles based on the validated multi-ancestry/East-Asian PRS model developed by Blechter et al. (2025) [15,16]. ETS exposure was classified as present or absent, yielding eight PRS– ETS strata. Relative risks (RRs) for PRS quartiles were derived from published estimates by combining epidermal growth factor receptor (EGFR) -positive and EGFR-negative odds ratios on the log scale using case-count weighting. ETS RRs were taken from meta-analyses and applied multiplicatively to PRS-specific baseline risks to generate combined PRS–ETS RRs. All formulas, weighting procedures, case counts, and RR values (point estimates and 95% CIs) are provided in Supplementary Methods and Supplementary Tables S1– S2.

### 2.4 Model structure

Eight independent state-transition microsimulation models were constructed, one for each PRS–ETS stratum. Each model was initialized at the stratum-specific optimal LDCT initiation age and simulated individuals annually until death or age 110. Incident lung adenocarcinomas entered directly at clinically detected stage, and individuals transitioned to post-treatment states or death according to stage-specific survival [10, 11, 13, 29].

Within each model, eight screening strategies were evaluated: annual LDCT, biennial LDCT, triennial LDCT, quadrennial LDCT, quinquennial LDCT, decennial LDCT, annual CXR, and no screening. Sensitivity and specificity values for LDCT and CXR were obtained from major screening trials and East Asian cohort studies [3, 11], and adherence rates were taken from observational studies and national surveys [11, 28]. Adherence to diagnostic follow-up after positive CXR was based on reasonable assumptions due to limited empirical evidence [Table 1].

**Table 1.**
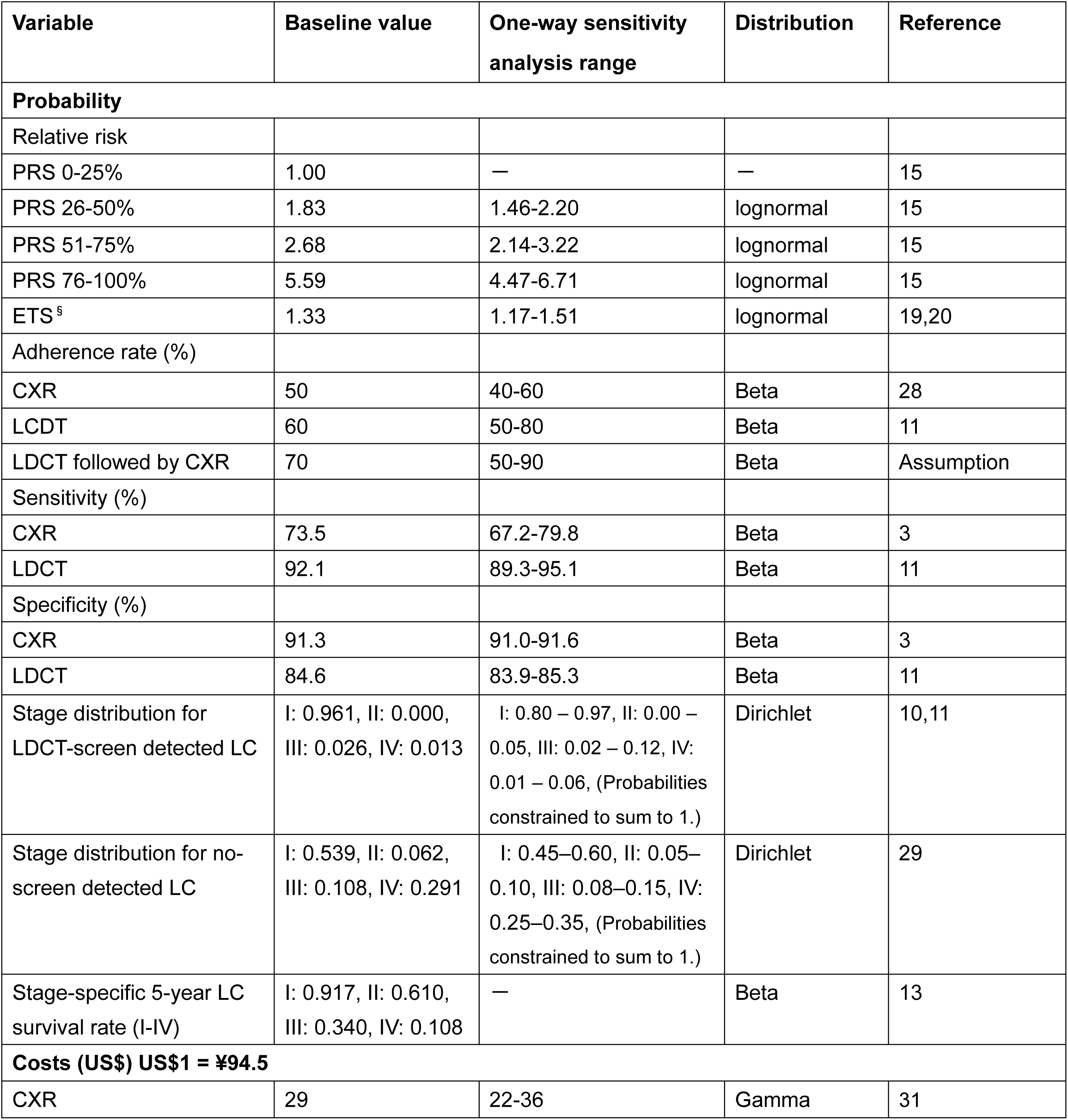
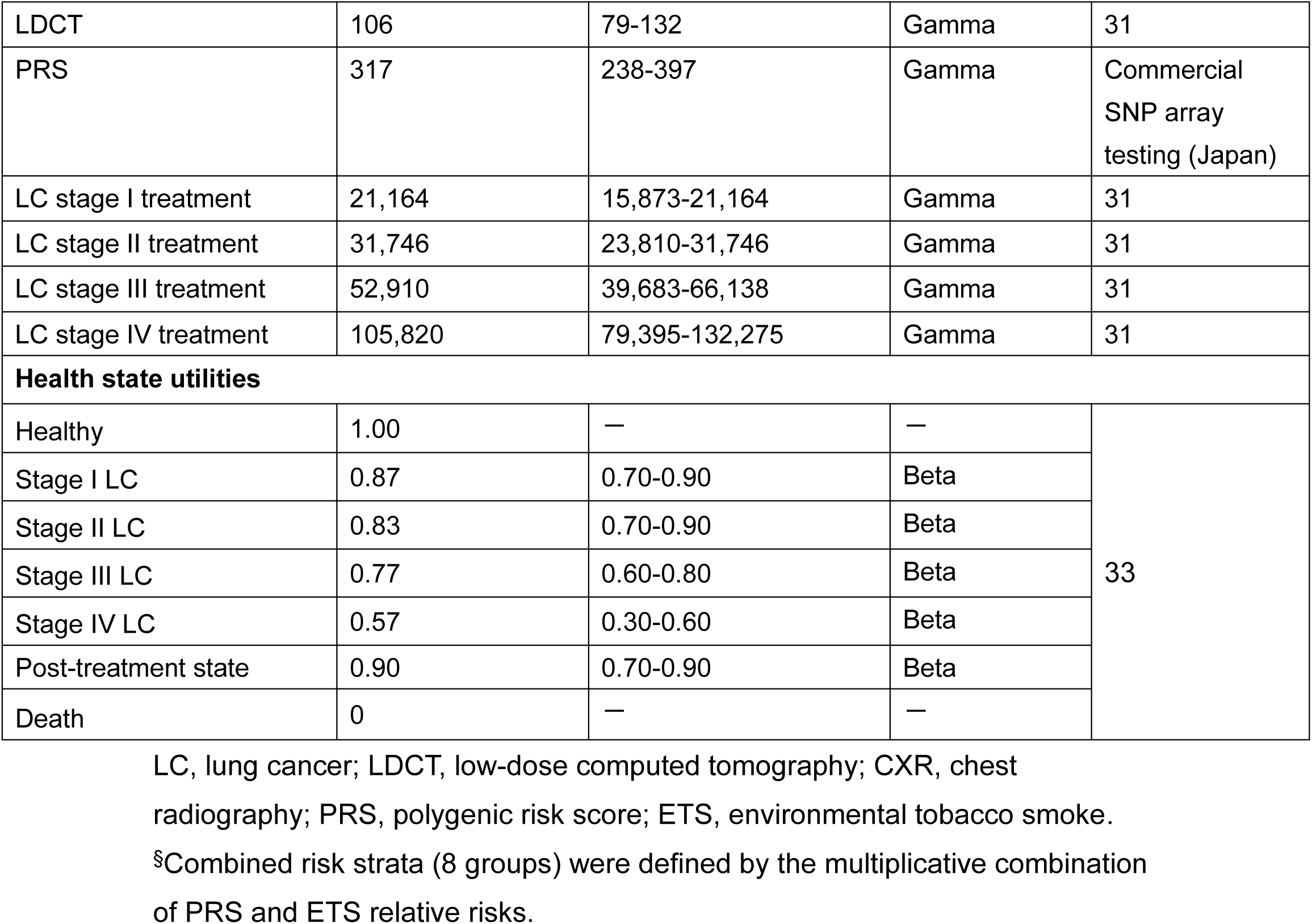
Baseline estimates for selected variables.

False positives were assumed to return to routine screening the following year and not proceed to immediate treatment, consistent with guideline-recommended diagnostic pathways [14, 23]. False negatives were modeled as cancers detected outside screening and were assigned the same stage distribution as unscreened cases based on national registry data [13, 29]. Overdiagnosis was not explicitly modeled because reliable estimates for non-progressive lung adenocarcinoma in East Asian never-smoker populations remain highly uncertain, and prior screening models have shown minimal impact of overdiagnosis assumptions on mortality and cost-effectiveness outcomes [4, 12].

### 2.5 Model inputs

Clinical probabilities and epidemiologic parameters were obtained from MEDLINE (2000–2026) searches and Japanese cancer statistics [30]. Baseline age-specific incidence of lung adenocarcinoma among never-smoking women was estimated using national cancer registry data, adenocarcinoma proportions, smoking prevalence, and published relative risks [9, 10, 28, 30].

Mixed-population incidence was decomposed into never-smoker–specific incidence using published relative risks [9].

Sensitivity and specificity for LDCT and CXR were obtained from a prospective cohort study in Taiwan and the National Lung Screening Trial [3, 11]. LDCT was assumed to shift the stage distribution toward earlier detection, with 96.1% of screen-detected cancers diagnosed at stage I based on Japanese observational data [10] (Table 1). Clinically detected cancers followed observed Japanese distributions (53.9% stage I) [29]. Stage distributions were modeled using Dirichlet distributions, and stage-specific survival was applied based on national survival statistics [13].

Direct medical costs were estimated from a healthcare-payer perspective. Screening costs were obtained from the Japanese fee schedule [31]. PRS testing cost (US$317) reflects the commercial SNP-array genomic test price used in the base case and is treated as an assumption; source and alternative values are reported in Table 1. Treatment costs were assigned according to Japanese lung cancer treatment guidelines [23]. All costs were converted to US dollars using the 2024 OECD purchasing power parity rate for Japan (1 US$ = ¥94.5) [32] and discounted at 3% annually. [26]

### 2.6 Health state utilities

The model included seven health states: healthy, lung cancer stages I–IV, post-treatment, and death (Figure 1). Health state utilities were obtained from published literature [33] and applied to estimate QALYs. All utilities were discounted at 3% annually in accordance with standard cost-effectiveness guidelines [26].

**Figure 1.**
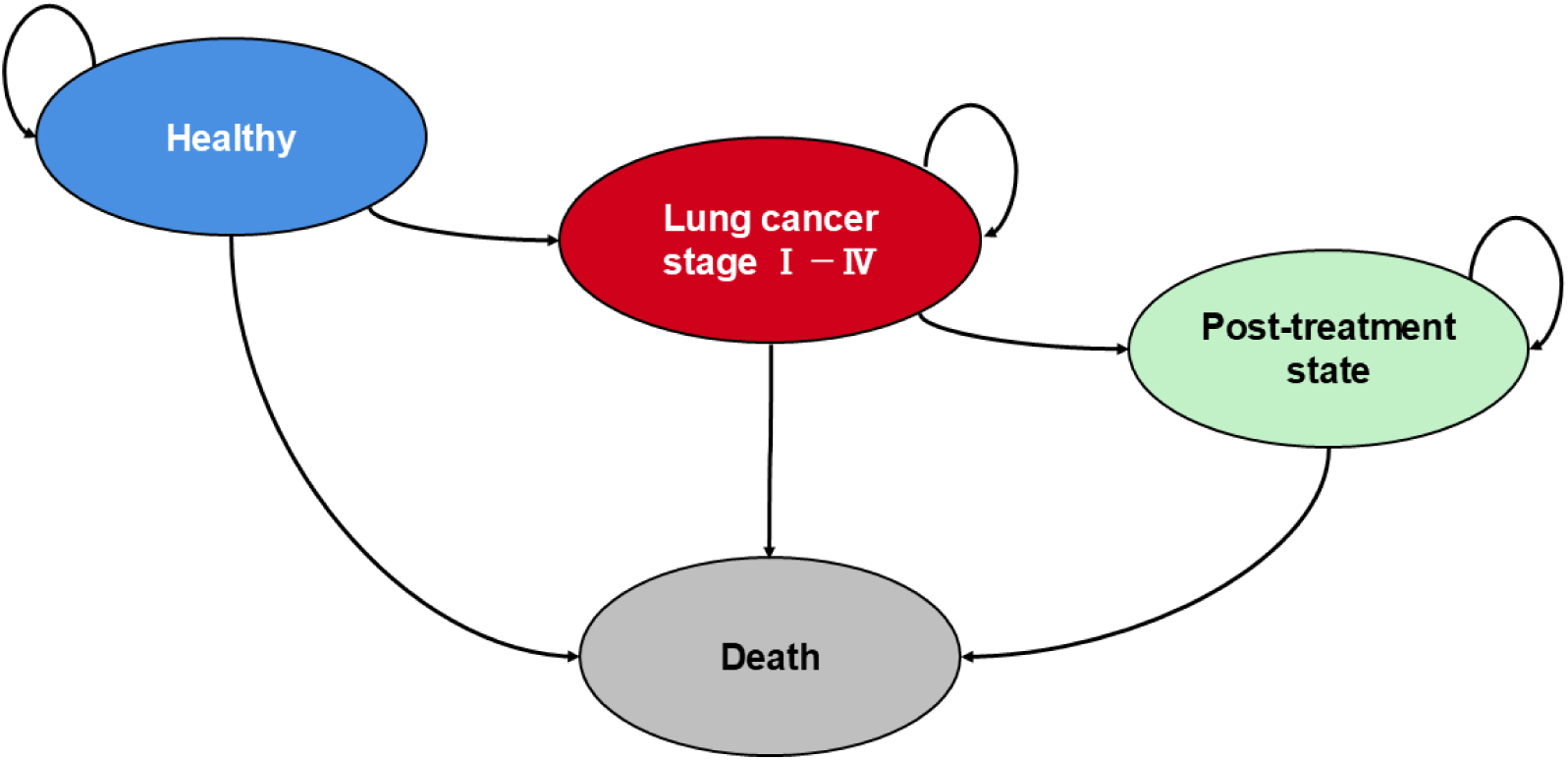
State transition structure of the lung cancer natural history model. This diagram illustrates the health states and possible transitions used in the lung cancer simulation model. Individuals begin in the healthy state, may progress to lung cancer stages I–IV, and subsequently transition to post-treatment or death. Arrows represent allowable transitions between states in the model.

### 2.7 Sensitivity analyses

One-way sensitivity analyses varied key parameters across ranges shown in Table 1. Probabilistic sensitivity analysis was performed using second-order Monte Carlo simulation with 10,000 iterations; parameter uncertainty was propagated to costs and QALYs and results were summarized as cost-effectiveness acceptability curves and incremental cost-effectiveness planes. Beta distributions were assigned to probabilities and utilities, gamma distributions to costs, log-normal distributions to PRS- and ETS-related relative risks, and Dirichlet distributions to stage distributions. Correlations between related parameters (e.g., stage proportions across stages) were preserved where applicable. A willingness-to-pay threshold (WTP) of US$50,000 per QALY gained was applied following national reference values [25].

### 2.8 Scenario analyses

Scenario analyses evaluated how optimal LDCT initiation ages shifted under conditions of unavoidable ETS exposure, reflecting structural vulnerability to environmental risk [19, 20]. Three scenarios were modeled: household ETS exposure, workplace ETS exposure, and combined household plus workplace ETS exposure.

Workplace ETS exposure was assigned a higher relative risk than household ETS [19, 20]. This assumption reflected Japan’s historically elevated indoor smoking levels in occupational, restaurant, and service-industry settings.

Although workplace smoking has declined following recent smoke-free legislation, residual ETS exposure persists in some environments, making historical exposure patterns relevant for lifetime risk estimation. Supporting evidence included national surveys showing elevated occupational ETS exposure in Japan [22], international pooled analyses indicating higher risks in high-exposure workplaces [20], and cohort data from Taiwan demonstrating increased lung cancer risk among never-smoking women exposed to workplace ETS [17].

ETS relative risks were calibrated to represent moderate to high exposure scenarios, informed by Japanese meta-analytic estimates and international pooled analyses [19, 20]. For each PRS–ETS stratum, optimal LDCT initiation ages were recalculated under these exposure conditions using the same 5-year screening age categories (40, 45, 50, and 55 years).

To evaluate robustness within each scenario, sensitivity analyses varied ETS relative risks by ±20% (Base: 1.06–1.60; Household: 1.24–1.86; Workplace: 1.42–2.14; Combined: 2.21–3.31). Continuous variation in ETS risks was translated into discrete screening recommendations by remapping each perturbed risk profile to the 5-year initiation age categories.

All analyses were conducted using TreeAge Pro 2026 (TreeAge Software, Williamstown, MA). This economic evaluation followed the CHEERS 2022 reporting guidelines [34].

### 2.9 Model validation

Face validity was established by ensuring consistency between model structure, natural history assumptions, clinical pathways, stage distributions, treatment patterns, and parameter choices with Japanese lung cancer screening and treatment guidelines [14, 23].

Internal validity was assessed by confirming that the model reproduced expected transitions, stage distributions, and survival patterns under screening and no-screening scenarios.

External validity was evaluated by comparing model-generated age-specific lung cancer incidence and stage distribution at diagnosis with published Japanese epidemiologic data [9, 10, 11, 13, 29, 30]. For lung cancer mortality, the model’s natural history reproduced the expected age-related increase observed in Japanese vital statistics, although absolute mortality levels were lower because the model represents never-smoking women. Model outputs closely matched observed patterns (Supplementary Table S3). Supplementary Materials provide supporting details for the validation analyses.

## 3. Results

### 3.1 Optimal initiation age for screening

Threshold analyses identified clear switching points in optimal LDCT initiation ages across PRS–ETS relative-risk levels. When the combined relative risk exceeded 7.11, annual LDCT beginning at age 40 years yielded higher NMB than initiation at age 45 years. At a relative risk above 2.40, initiation at age 45 years became more cost-effective than starting at age 50 years. Similarly, when relative risk surpassed 1.44, initiation at age 50 years outperformed initiation at age 55 years. These thresholds demonstrate that higher PRS–ETS risk profiles favor earlier LDCT initiation (Supplementary Table S4).

Optimal initiation ages for annual LDCT screening varied substantially across PRS–ETS strata. Initiation ages ranged from 40 to 55 years: 55 years for PRS1 and PRS1+ETS, 50 years for PRS2, 45 years for PRS3, PRS4, PRS2+ETS, and PRS3+ETS, and 40 years for PRS4+ETS (Figure 2, Supplementary Table S5). These patterns indicate meaningful heterogeneity in optimal screening timing driven by underlying genetic and environmental risk.

**Figure 2.**
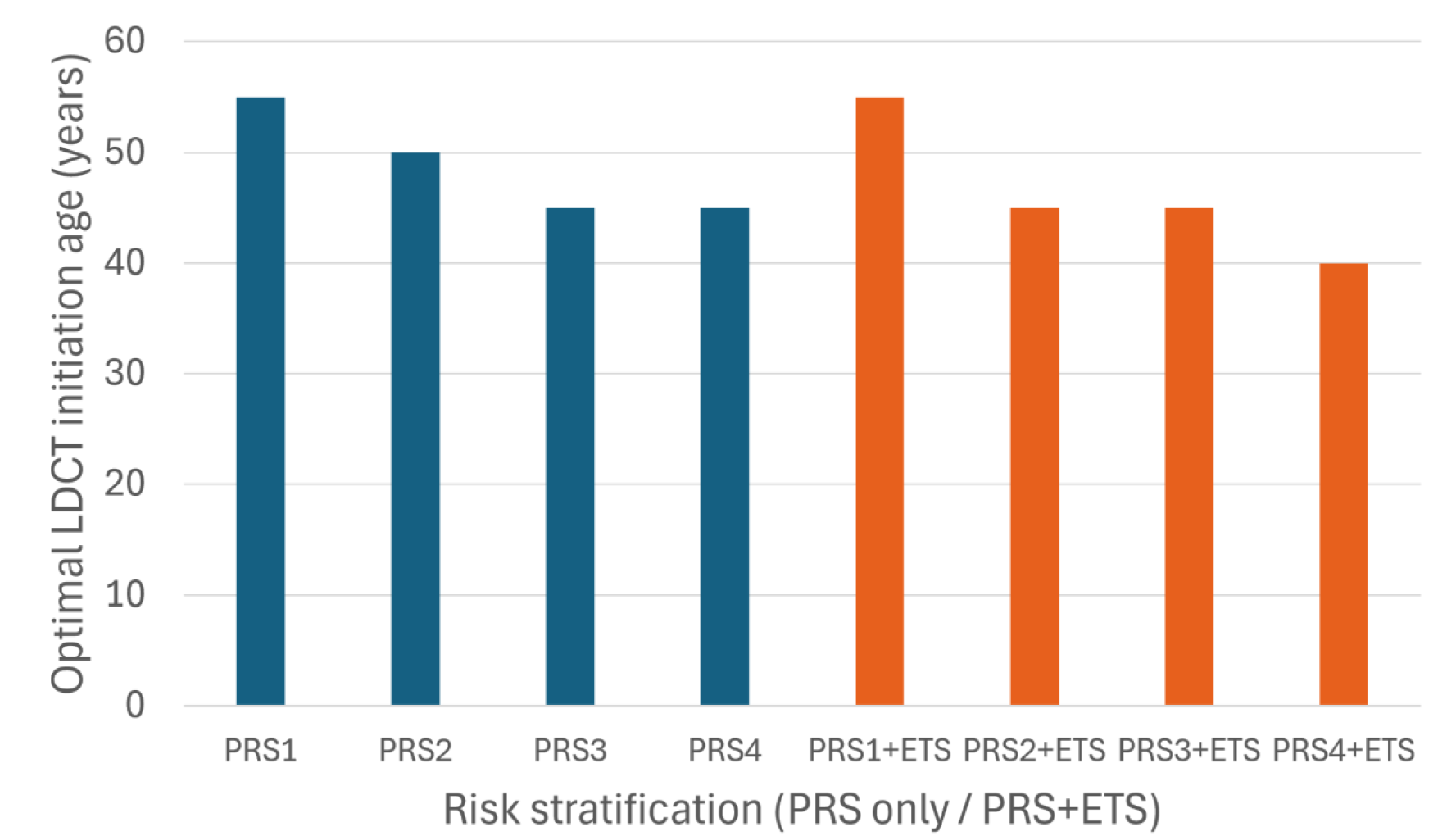
Optimal LDCT initiation age by PRS and ETS stratification. This figure shows the model-estimated optimal age to initiate LDCT screening under different risk stratification strategies. Blue bars represent initiation ages based on PRS alone, while orange bars incorporate both PRS and ETS. Adding ETS consistently shifts the optimal initiation age earlier across all PRS groups. LDCT, low-dose computed tomography; PRS, polygenic risk score; ETS, environmental tobacco smoke.

### 3.2 Base case analysis

Across all PRS–ETS strata, annual LDCT screening was the most cost-effective strategy (Table 2, Figure 3, Supplementary Figure S1, Supplementary Tables S6-S13). In the lowest-risk stratum (PRS1), annual LDCT yielded an ICER of US$40,471 per QALY compared with annual CXR, with ICERs declining progressively in higher-risk strata (Figure 4A). In the highest-risk groups (PRS4 and PRS4+ETS), annual LDCT was dominant and cost saving relative to both CXR and no screening. PRS4+ETS yielded the highest NMB among all strategies (Figure 4B).

**Figure 3.**
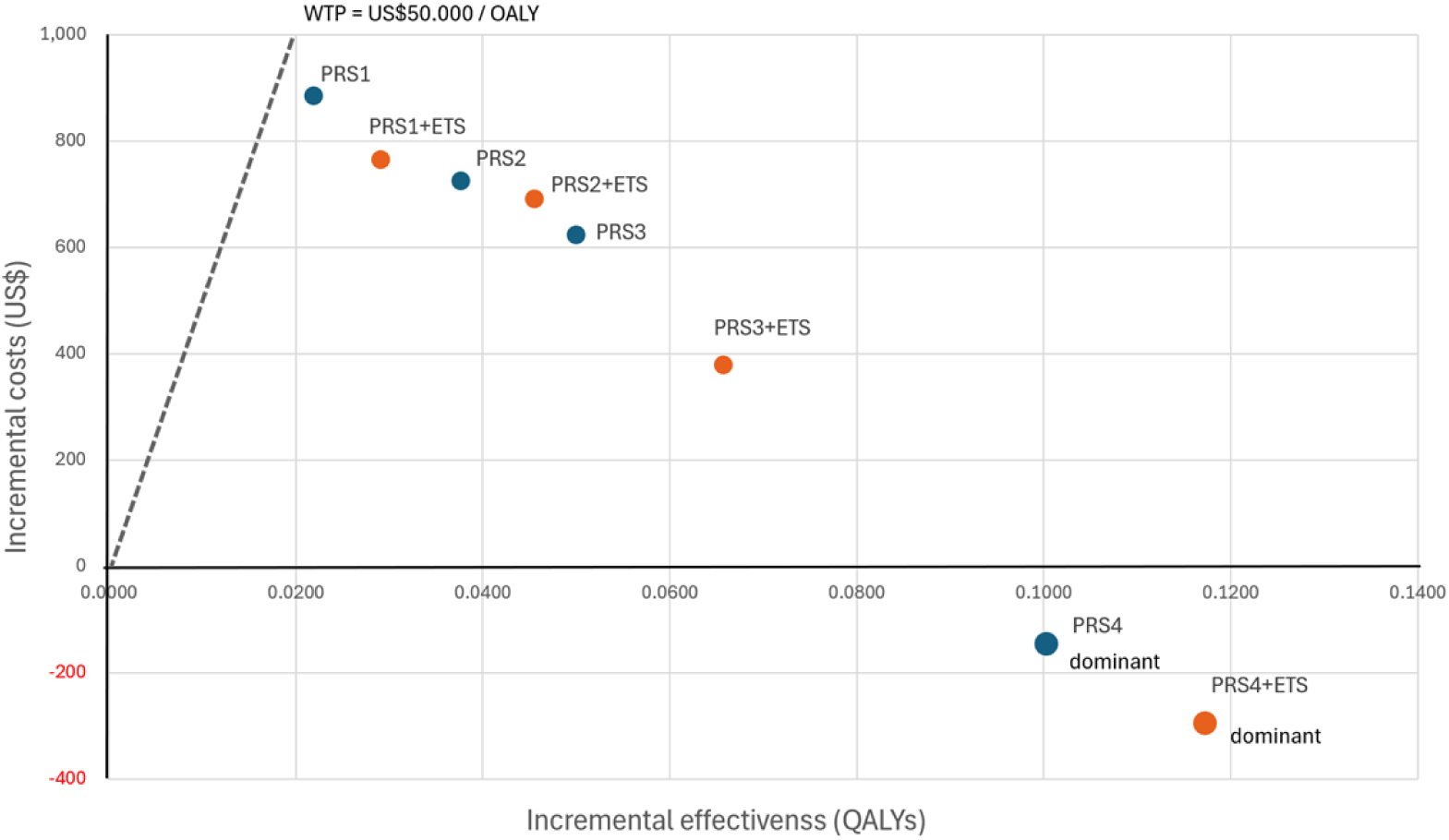
Cost-effectiveness plane of annual LDCT strategies across PRS and PRS+ETS risk strata. Incremental costs and incremental QALYs are shown relative to the current screening policy. Strategies in the lower-right quadrant (e.g., PRS4 and PRS4+ETS) are dominant, providing greater effectiveness at lower cost. PRS1– PRS3 and their ETS combinations yield positive incremental costs with varying gains in QALYs. The willingness-to-pay (WTP) threshold was set at US$50,000 per QALY. Strategies below this line are considered cost-effective relative to the current policy. LDCT, low-dose computed tomography; PRS, polygenic risk score; ETS, environmental tobacco smoke.

**Figure 4.**
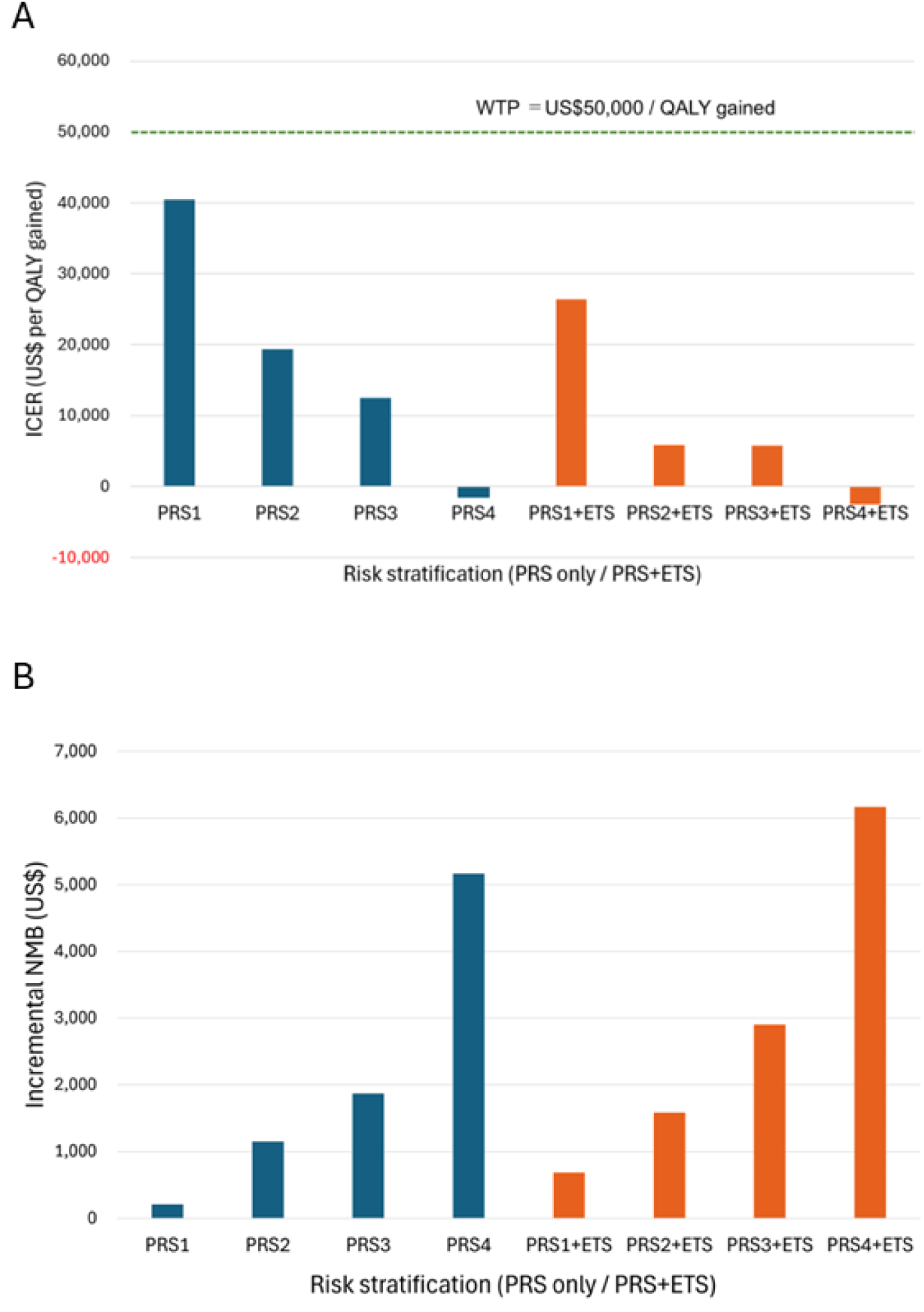
Cost-effectiveness outcomes of annual LDCT strategies across PRS and PRS+ETS risk strata. A. Incremental cost-effectiveness ratios (ICERs) ICERs are calculated relative to annual CXR. The horizontal line indicates the willingness-to-pay (WTP) threshold of US$50,000 per QALY gained. PRS4 and PRS4+ETS are dominant strategies, providing greater effectiveness at lower cost. B. Incremental net monetary benefits (NMBs) NMB is calculated using a WTP threshold of US$50,000 per QALY gained. Higher NMB values indicate greater economic benefit relative to annual CXR. PRS4+ETS yields the highest NMB among all strategies. LDCT, low-dose computed tomography; CXR, chest radiography; PRS, polygenic risk score; ETS, environmental tobacco smoke.

**Table 2.** Cost-Effectiveness outcomes of annual LDCT screening across PRS and PRS+ETS risk strata.

| Risk stratum | Incremental Costs (US\$) | Incremental QALYs | ICERs (US\$ per QALY gained) | Incremental NMBs (US\$) |
| --- | --- | --- | --- | --- |
| PRS1 | 886 | 0.0219 | 40,471 | 209 |
| PRS2 | 727 | 0.0376 | 19,334 | 1,153 |
| PRS3 | 624 | 0.0499 | 12,508 | 1,871 |
| PRS4 | -147 | 0.1004 | -1,469 | 5,167 |
| PRS1+ETS | 766 | 0.0290 | 26,362 | 687 |
| PRS2+ETS | 692 | 0.0455 | 5,849 | 1,584 |
| PRS3+ETS | 380 | 0.0657 | 5,779 | 2,906 |
| PRS4+ETS | -297 | 0.1174 | -2,526 | 6,165 |

LDCT, low-dose computed tomography; PRS, polygenic risk score; ETS, environmental tobacco smoke; QALY, quality-adjusted life year; ICER, incremental cost-effectiveness ratio; NMB, net monetary benefit.

### 3.3 One-way sensitivity analysis

Deterministic one-way sensitivity analyses demonstrated high robustness of the cost-effectiveness results. Four parameters—discount rate, LDCT stage I detection probability, LDCT adherence, and adherence to diagnostic follow-up after positive CXR—produced modest shifts in ICERs in the lowest-risk stratum (PRS1) (Supplementary Table S14, Figure S2). None altered the preferred strategy. All higher-risk strata (PRS2–PRS4 and all PRS+ETS groups) showed no meaningful variation in ICERs under any parameter change.

### 3.4 Probabilistic sensitivity analysis

Probabilistic sensitivity analysis confirmed the stability of the base-case results. Across 10,000 Monte Carlo simulations, the probability that annual LDCT was cost effective at a WTP threshold of US$50,000 per QALY increased progressively with genetic and environmental risk. In strata without ETS exposure, annual LDCT was cost effective in 45.1%–68.8% of simulations (PRS1–PRS4) (Figure 5). ETS exposure further increased these probabilities to 50.6%–70.9% across PRS1+ETS to PRS4+ETS.

**Figure 5.**
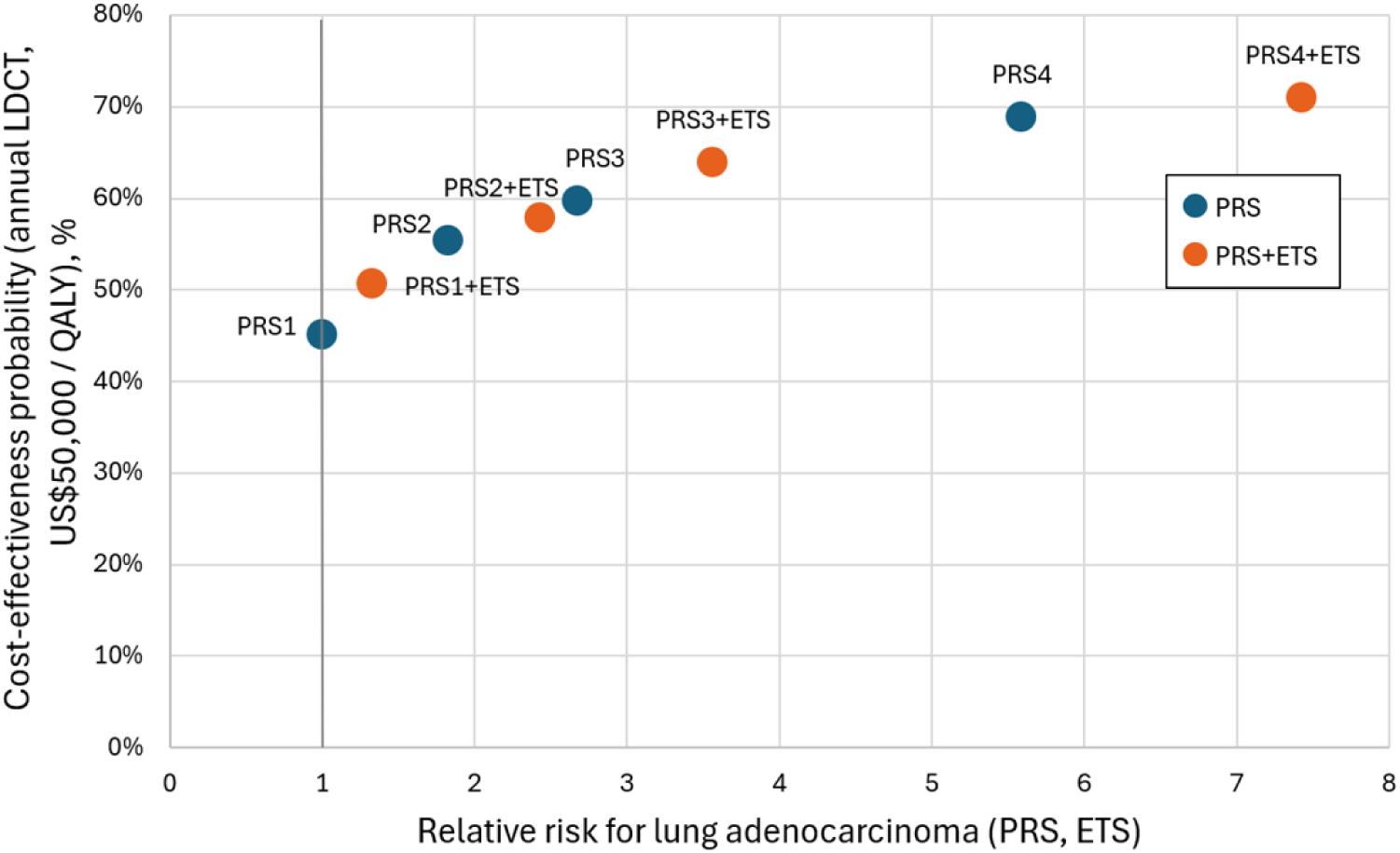
Cost-effectiveness probabilities for annual LDCT strategies at a WTP threshold of US$50,000 per QALY gained across PRS and PRS+ETS risk strata. LDCT, low-dose computed tomography; PRS, polygenic risk score; ETS, environmental tobacco smoke.

### 3.5 Scenario analyses

Scenario analyses showed that unavoidable ETS exposure shifted optimal LDCT initiation ages earlier across PRS–ETS strata. Household or workplace ETS exposure advanced initiation ages from the standard 50–45-year range to as early as 40 years in higher-risk strata (Table 3). Combined household and workplace exposure produced the largest downward shifts, decreasing initiation ages to 45 years in PRS1+ETS and to 40 years in PRS4+ETS. These findings indicate a measurable early bias in optimal screening timing driven by involuntary environmental exposure.

**Table 3.** Downward Shift in Optimal LDCT Initiation Ages Across PRS–ETS Strata by ETS Exposure Setting.

| Risk Stratum | Base (ETS RR=1.33) | Household (ETS RR=1.55) | Workplace (ETS RR=1.78) | Combined Household + Workplace (ETS RR=2.76) |
| --- | --- | --- | --- | --- |
| PRS1+ETS | 55 | 50 | 50 | 45 |
| PRS2+ETS | 45 | 45 | 45 | 45 |
| PRS3+ETS | 45 | 45 | 45 | 40 |
| PRS4+ETS | 40 | 40 | 40 | 40 |

LDCT, low-dose computed tomography; PRS, polygenic risk score; ETS, environmental tobacco smoke; RR, relative rate.

Sensitivity analyses varying ETS relative risks by ±20% resulted in at most a single 5-year shift in optimal initiation ages across all strata (Supplementary Table S15). Larger shifts were observed in lower-risk strata (PRS1–PRS2), where small changes in risk estimates naturally influence borderline screening decisions. In contrast, initiation ages in higher-risk strata (PRS3–PRS4) remained largely unchanged, indicating strong robustness of the primary conclusions. These shifts reflect the disproportionate burden of involuntary ETS exposure historically experienced by women, highlighting a structural inequity in lung cancer risk.

### 3.6 Cumulative lifetime costs and health outcomes

Annual LDCT screening increased lifetime QALYs and shifted diagnoses toward earlier stages, reducing late-stage disease and lung adenocarcinoma mortality. Deaths averted increased steadily with higher polygenic risk, and ETS exposure further amplified these gains (Figure 6). The largest reductions were observed in PRS4 and PRS4+ETS, with the latter approaching nearly 3,000 deaths averted.

**Figure 6.**
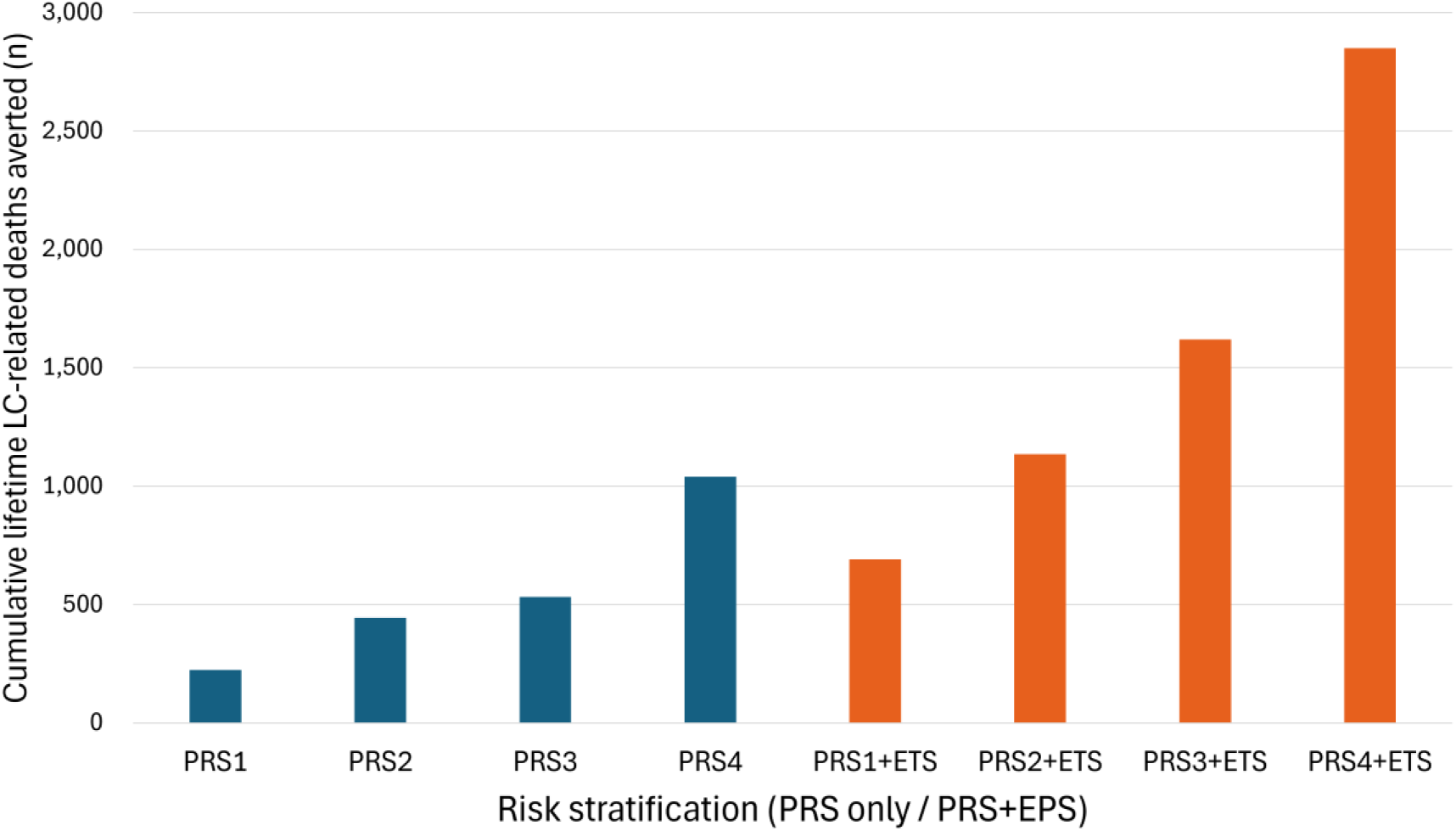
Cumulative lifetime lung adenocarcinoma-related deaths averted by annual LDCT strategy compared with annual CXR strategy across PRS and PRS+ETS risk strata. Higher-risk strata (such as PRS4 and PRS4+ETS) show substantially greater cumulative lifetime lung adenocarcinoma–related deaths averted than lower-risk strata. LDCT, low-dose computed tomography; CXR, chest radiography; PRS, polygenic risk score; ETS, environmental tobacco smoke.

At the population level, annual LDCT substantially increased stage I detection and reduced stages II–IV (Supplementary Table S16). Over a lifetime, annual LDCT averted 8,534 lung adenocarcinoma-related deaths compared with annual CXR and 14,940 deaths compared with no screening.

## Discussion

### Key findings

Annual LDCT screening for never-smoking women was the most cost-effective strategy, yielding the largest reductions in lung adenocarcinoma mortality and substantially lowering the proportion of late-stage diagnoses.

To our knowledge, this is the first study to evaluate risk-stratified LDCT screening strategies for never-smoking women using combined PRS–ETS risk, and to identify the optimal initiation age for precision screening.

PRS stratification identified women who would benefit from earlier screening initiation, with optimal ages decreasing to 40 years in the highest-risk stratum. Adding ETS exposure revealed a substantial, involuntary risk burden, often concentrated among socially vulnerable women. This burden further shifts optimal initiation ages under combined exposure scenarios. Together, these findings show that precision screening with risk-stratified initiation ages can support earlier, potentially curable detection of lung adenocarcinoma among never-smoking women while providing a more equitable framework for prevention.

### Interpretation

These findings demonstrate that risk-stratified LDCT screening can correct a major misalignment between the current annual CXR-only approach for never-smoking women and the need for annual LDCT screening among the populations at highest risk. Prior studies have noted that never-smoking Asian women often develop lung adenocarcinoma despite minimal personal tobacco exposure, yet no screening framework has incorporated the combined effects of genetic susceptibility and involuntary ETS exposure. Our results show that these factors jointly produce lifetime risks approaching those of moderate smokers, offering a biologically and environmentally grounded rationale for earlier LDCT initiation. Importantly, annual LDCT markedly reduced late-stage diagnoses in our models, suggesting that precision screening could counter the persistent pattern of advanced-stage presentation in this population. The stability of initiation ages in higher-risk strata indicates that recommendations for these groups remain robust even under uncertainty in ETS risk estimates, while modest shifts in lower-risk strata reflect the expected sensitivity of borderline screening decisions. Overall, integrating PRS–ETS risk information provides a more accurate framework for identifying never-smoking women at greatest risk of developing potentially curable lung adenocarcinoma.

Beyond clinical effectiveness, our findings highlight a critical equity dimension. Women with socially vulnerable backgrounds, who are disproportionately exposed to involuntary ETS in homes, workplaces, and public environments, accumulate lung cancer risk earlier in life despite never smoking. This structural disadvantage shifts optimal LDCT initiation toward younger ages in precisely those groups least protected by current screening policy. Integrating genetic susceptibility with involuntary environmental exposure therefore provides not only a more accurate risk-stratification framework but also an ethically grounded rationale for correcting inequities in lung cancer prevention among never-smoking women.

### Comparison with existing literature

Randomized LDCT trials such as NLST and NELSON [3, 4] enrolled only heavy smokers, leaving never-smoking women entirely unrepresented. Existing risk prediction models rarely incorporate PRS and almost never include ETS exposure, leaving major sources of risk among never-smoking women unaccounted for. Under the 2025 Japanese lung cancer screening guideline [14], annual LDCT is recommended only for heavy smokers aged 50–74 years, and LDCT is not advised for never smokers because of insufficient evidence.

Our results show that never-smoking women, as a group already at elevated risk for lung adenocarcinoma, can reach lifetime risk levels that overlap with, and in some cases exceed, those of guideline-eligible smokers, particularly when both PRS and ETS exposure are high. These findings suggest that current eligibility criteria may be overly restrictive.

Most prior epidemiologic models treat screening as a single-test process, representing false positives and false negatives as simple misclassification probabilities without modeling downstream diagnostic pathways. In contrast, our LDCT-based screening model reflects the hierarchical diagnostic structure used in clinical practice, in which abnormal CXR or LDCT findings trigger further imaging, surveillance, or clinical evaluation. Consequently, false positives typically return to routine screening without unnecessary treatment, while false negatives proceed to symptomatic detection with stage distributions similar to unscreened cases. This clinically grounded representation models repeated LDCT screening at intervals of 1, 2, 3, 4, 5, and 10 years and allows false positives to return to routine screening and false negatives to enter symptomatic detection. It differs fundamentally from conventional single-test abstractions.

Because simpler frameworks do not capture repeated screening or these basic downstream processes, their expected outcomes naturally diverge from those produced by our model.

### Policy implications

These findings indicate that LDCT eligibility criteria in Japan and other Asian countries should be reconsidered, particularly for never-smoking women in whom peripheral adenocarcinoma predominates and LDCT provides substantially higher sensitivity than CXR. CXR-based programs therefore offer limited effectiveness for this population. A risk-stratified approach to initiation age may offer a practical and equitable framework, enabling women with elevated PRS or ETS to begin screening at 40–45 years rather than 50–55.

Because ETS exposure reflects structural inequities shaped by workplace and socioeconomic conditions and accelerates lifetime risk accumulation, earlier LDCT initiation for exposed women could function as an equity-enhancing intervention. Strengthening smoke-free environment policies, including enforcement of workplace protections, would further reduce ETS exposure and complement risk-based screening strategies. However, uncertainties regarding PRS calibration in Asian populations, ETS measurement, and health-system capacity for LDCT expansion, including management of false positives and overdiagnosis, warrant a phased implementation. Pilot risk-stratified LDCT programs with embedded evaluation of effectiveness, harms, and cost-effectiveness, together with prospective validation of PRS and ETS metrics, would support evidence-based guideline refinement. Incorporating PRS and ETS into eligibility criteria through validated, staged adoption could reduce mortality among populations underserved by smoking-based guidelines and align screening policy with the contemporary epidemiology of lung cancer in Asia.

### Strengths

This study is the first to integrate PRS with ETS exposure to generate lifetime lung cancer risk trajectories tailored to never-smoking Asian women. Linking these trajectories to an individual-level microsimulation enables systematic evaluation of risk-stratified initiation ages and screening intervals across diverse genetic and environmental profiles. This framework establishes an evidence base for future updates to lung cancer screening guidelines and supports development of equitable, risk-based early detection strategies for populations historically overlooked by smoking-based eligibility criteria.

Use of nationally representative Japanese incidence, cost, and diagnostic data enhances internal validity, and the model reflects real-world diagnostic pathways, including widespread CXR use, allowing accurate comparison of screening modalities. Evaluating multiple initiation ages across eight risk strata yields granular evidence for precision screening, while comprehensive cost and QALY estimation strengthens robustness.

In addition, we developed a threshold-based extension of one-way sensitivity analysis to identify the relative-risk levels at which the optimal LDCT initiation age switches between discrete strategies. By running parallel Markov models initiating annual LDCT at 40, 45, 50, 55, 60, and 65 years and varying the combined PRS–ETS relative risk from 1 to 8, we derived the relative-risk thresholds at which net monetary benefit favored earlier versus later initiation. This methodological innovation has not been previously applied in lung cancer screening models and provides a practical framework for determining optimal initiation ages when strategies are defined in discrete intervals.

Overdiagnosis was modeled using conservative assumptions aligned with Japanese practice, in which indolent lesions are typically managed through surveillance. Incorporating ETS as a structural exposure adds an equity-focused dimension rarely included in lung cancer screening models. Parameterization was informed by published evidence on adenocarcinoma natural history in East Asian never-smoking women, which enhances biological plausibility without relying on unobservable tumor-growth parameters. This study employs an incidence-based modelling framework rather than a natural-history-based model. This approach minimizes reliance on unobservable tumor-growth parameters, which remain highly uncertain for adenocarcinoma in never-smoking Asian women, and enables transparent integration of PRS and ETS-derived lifetime risk trajectories. By grounding all transitions in empirically observed incidence patterns, the model provides a robust and reproducible structure suitable for evaluating risk-stratified LDCT strategies in populations where natural-history evidence is limited. Integrating mortality reduction, overdiagnosis, costs, and QALYs within a unified framework provides a holistic assessment of LDCT value across heterogeneous risk groups. Diagnostic misclassification was explicitly incorporated: false negatives were modeled as missed detections that subsequently entered symptomatic clinical pathways within the same year, while false positives transitioned into annual surveillance consistent with Japanese LDCT practice. This approach ensures that sensitivity, specificity, and downstream consequences are coherently reflected in costs, QALYs, and overdiagnosis estimates.

### Limitations

Radiogenic cancer risk from repeated LDCT was not included [35], although prior analyses suggest that lifetime excess risk is too small to influence cost effectiveness and substantially increases model complexity [24]. ETS exposure was represented using cross-sectional estimates [19, 20], and more detailed longitudinal measurements could improve risk-stratification accuracy. Other risk factors, such as indoor air pollution, were not modeled because relevant data are limited. Cost inputs were based on Japanese healthcare prices [31], and applying this model to other Asian countries would require country-specific cost structures.

### Future directions

Future work should evaluate AI-assisted LDCT interpretation to reduce inter-reader variability and improve diagnostic reliability. Improving adherence will be essential for maximizing population-level benefits, and potential strategies include public awareness campaigns, reminder systems, and community-based outreach. Further research should examine the feasibility of integrating PRS into routine screening workflows, including cost, acceptability, and operational requirements. Refining ETS exposure metrics, particularly longitudinal and workplace-specific assessments, may enhance risk-stratification accuracy. Establishing the infrastructure required for PRS- and ETS-based risk-stratified LDCT screening will be essential for adoption across diverse health systems in Asia. This includes adequate CT capacity, trained radiologists, surveillance systems, and sustainable financing arrangements.

Risk stratified optimization of LDCT initiation age based on combined genetic and environmental risk profiles may provide a foundational framework for future personalized lung cancer screening. As PRS and ETS assessment become more widely available, tailoring screening start ages to individual risk rather than applying uniform age thresholds could enable safer, more efficient, and more equitable screening strategies that ultimately improve outcomes for never-smoking women.

Emerging technologies may further complement LDCT-based risk stratification in the future. Liquid biopsy approaches, including circulating tumor DNA assays, show promise for detecting radiographically occult early-stage adenocarcinoma [36]. Breath-based volatile organic compound profiling, protein-based multi-cancer early detection tests, and ultra-low-dose CT also warrant evaluation as potential adjuncts for safer and more sensitive screening in never-smoking women.

## Conclusion

Tailoring LDCT initiation age across PRS–ETS risk groups maximizes the mortality reduction achievable with cost-effective annual LDCT screening. PRS– ETS risk-stratified annual LDCT screening is substantially more cost-effective than annual CXR, and its greatest value emerges when initiation age is tailored to underlying risk, varying from 40 to 55 years. Optimal LDCT initiation age shifts systematically with underlying PRS–ETS risk, revealing that screening should begin earlier for women whose lifetime adenocarcinoma risk accumulates more rapidly. This dynamic alignment reduces the likelihood of late-stage detection that currently drives poor outcomes in this population.

These findings expose a fundamental limitation of global lung cancer guidelines that rely exclusively on smoking history and underscore the epidemiologic and scientific rationale for LDCT implementation in Asian never-smoking women, whose rising adenocarcinoma burden demands screening frameworks that reflect the evolving disease profile.

## Data Availability

All data used in this study are publicly available.

