## Supplementary Material for "Optimal LDCT screening for never-smoking Asian women using integrated polygenic and environmental risk: a microsimulation modelling study"

### Supplementary materials

#### Table of Contents


**Supplementary Methods**

**Supplementary Table S1.** EGFR-positive and EGFR-negative odds ratios used for PRS integration

**Supplementary Table S2.** Combined PRS–ETS relative risks used in the microsimulation model

**Supplementary Table S3.** Age-specific lung cancer incidence: observed vs model (never-smoking women)

**Supplementary Table S4.** Threshold values from one-way sensitivity analyses for strategy switching points in initiation age for annual LDCT in the PRS–ETS model

**Supplementary Table S5.** Optimal screening initiation age by PRS and ETS risk strata

**Supplementary Table S6.** Base-case results for the PRS1 stratum

**Supplementary Table S7.** Base-case results for the PRS2 stratum

**Supplementary Table S8.** Base-case results for the PRS3 stratum

**Supplementary Table S9.** Base-case results for the PRS4 stratum

**Supplementary Table S10.** Base-case results for the PRS1+ETS stratum

**Supplementary Table S11.** Base-case results for the PRS2+ETS stratum

**Supplementary Table S12.** Base-case results for the PRS3+ETS stratum

**Supplementary Table S13.** Base-case results for the PRS4+ETS stratum

**Supplementary Table S14.** Threshold Values Identified in One-Way Sensitivity Analysis for the PRS1 Stratum

**Supplementary Table S15.** Optimal LDCT initiation age under ETS relative risk scenarios (−20%, baseline, +20%)

**Supplementary Table S16.** Population-level lifetime costs and health outcomes for annual LDCT versus annual CXR and versus no screening

**Supplementary Figure S1.** Efficiency frontier for LDCT and CXR across EPS status and PRS levels

**Supplementary Figure S2.** Tornado diagram for PRS1 stratum

#### Supplementary Methods.

##### Model Overview

These supplementary materials provide detailed descriptions of PRS–ETS integration, model parameterization, validation procedures, and full cost-effectiveness results across all risk strata.

##### 1. Polygenic Risk Score (PRS)

The PRS used in this study was derived from the multi-ancestry/East-Asian lung adenocarcinoma PRS model developed by Blechter et al. (2025). The model provides odds ratios (ORs) for lung adenocarcinoma among epidermal growth factor receptor (EGFR)-positive and EGFR-negative cases across PRS quartiles. PRS values were categorized into quartiles (0–25%, 26–50%, 51–75%, 76–100%) based on the distribution reported in the original study.

All EGFR-specific ORs and sample counts used for PRS integration are listed in Supplementary Table S1.

##### 2. Integration of EGFR-positive and EGFR-negative Odds Ratios

Because the PRS model reports separate ORs for EGFR-positive and EGFR-negative lung adenocarcinoma, we combined these estimates into a single PRS-specific relative risk applicable to the overall never-smoker population.

Let $OR_{EGFR+}$ and $OR_{EGFR-}$ denote the multinomial ORs for EGFR-positive and EGFR-negative disease, and let $n_{EGFR+}$ and $n_{EGFR-}$ denote the corresponding case counts.

The combined log-scale estimate was:

$$\log(OR_{\text{combined}}) = \frac{n_{EGFR+} \cdot \log(OR_{EGFR+}) + n_{EGFR-} \cdot \log(OR_{EGFR-})}{n_{EGFR+} + n_{EGFR-}}$$

The final combined OR was obtained by exponentiation:

$$OR_{\text{combined}} = \exp\left(\log(OR_{\text{combined}})\right)$$

The resulting PRS-specific relative risks were:

- Quartile 1: 1.00
- Quartile 2: 1.83
- Quartile 3: 2.68
- Quartile 4: 5.59

These values are shown in Supplementary Table S1 and were used as baseline relative risks in the microsimulation model.

**3. Environmental Tobacco Smoke (ETS)**

Relative risks for environmental tobacco smoke (ETS) exposure were obtained from the meta-analysis by Ni et al. (2018), which synthesized evidence across household, workplace, and community exposure studies among women.

$$RR_{\text{ETS}} = 1.33\ (95\%\ CI: 1.17\text{–}1.51)$$

This estimate was applied uniformly across PRS strata.

**4. Combined PRS–ETS Relative Risks**

We assumed multiplicative joint effects of PRS and ETS exposure:

$$RR_{\text{PRS–ETS}} = RR_{\text{PRS}} \times RR_{\text{ETS}}$$

This produced eight strata (four PRS quartiles × ETS present/absent). All combined relative risks are listed in Supplementary Table S2.

**5. Screening Test Characteristics**

Sensitivity and specificity values for chest radiography (CXR) and low-dose computed tomography (LDCT) were obtained from published Japanese and international screening studies. Stage distributions for screen-detected and non-screen-detected lung cancer were taken from registry and hospital-based data. Stage-specific survival rates were derived from Japanese cancer statistics and clinical outcome studies.

All screening performance parameters, costs, and utilities used in the model are summarized in Table 1.

**6. Model Inputs**

The model was incidence-based and did not rely on explicit natural history transition rates. Age-specific incidence, stage distribution at diagnosis, and lung cancer mortality rates for never-smoker women were taken directly from Japanese registry and cancer statistics data (see Table 1)

**7. Parameterization of Probability Distributions**

Probability and utility parameters were modeled using beta distributions.
Beta distribution parameters (α, β) were derived from reported means (μ) and standard errors (SE) using:

$\alpha = \mu \times [(\mu \times (1 - \mu) / SE^2) - 1]$

$\beta = (1 - \mu) \times [(\mu \times (1 - \mu) / SE^2) - 1]$.

Cost parameters were modeled using gamma distributions.
Gamma distribution parameters were calculated from means (μ) and standard errors (SE) as:

$\text{shape} = (\mu / SE)^2$

$\text{scale} = SE^2 / \mu$.

Relative risks associated with PRS and ETS were modeled using log-normal distributions.
Log-normal parameters (μ_ln, σ_ln) were derived from reported point estimates and 95% confidence intervals (CI):

$\mu\_ln = \ln(RR)$

$\sigma\_ln = [\ln(\text{upper CI}) - \ln(\text{lower CI})] / (2 \times 1.96)$.

Stage distributions were modeled using Dirichlet distributions.

Dirichlet parameters ($\alpha_1$, $\alpha_2$, …, $\alpha_k$) were assigned proportional to observed stage frequencies, scaled by the sample size of the underlying data source.

All baseline values, one-way sensitivity analysis ranges, assigned distributions, and references are listed in Table 1.

**8. PSA Convergence Diagnostics**

Probabilistic sensitivity analysis (PSA) was conducted using 10,000 second-order Monte Carlo iterations. Convergence was assessed by examining the stability of mean costs, QALYs, and incremental cost-effectiveness ratios across increasing numbers of iterations. Cost-effectiveness acceptability curves (CEACs) and incremental cost-effectiveness planes showed smooth and stable patterns without evidence of divergence, indicating adequate convergence of the PSA. Relevant PSA outputs are presented in Figure 5.

**9. Model Validation Details**

**9-1. External validation: Age-specific incidence comparison**

Table S3 compares model-generated age-specific lung cancer incidence with observed incidence from Japanese Cancer Statistics. Although absolute rates differ because the model represents never-smoking women, the age-specific pattern of increasing incidence with age was consistent between model outputs and observed data.

**9-2. External validation: Stage distribution at diagnosis**

The model used stage distribution at diagnosis for lung cancer based on published Japanese data from Kakinuma et al. (2020). Stage I–IV proportions in the model were aligned with the observed distribution reported in this study, ensuring consistency between model assumptions and empirical Japanese registry data.

**9-3. External validation: Lung cancer mortality**

The component of the model was calibrated so that lung cancer mortality increases with age, consistent with the pattern observed in Japanese cancer statistics. Because the model represents never-smoking women, absolute mortality levels are lower than population-level mortality, but the age-related pattern is aligned with empirical data.

##### 9-4. Internal validation checks

The following internal consistency checks were performed:

- Cohort counts were preserved across all branches.
- Event ordering followed clinical logic (incidence → detection → treatment → survival).
- Screening produced expected stage shifts (higher early-stage detection).
- Survival curves under screening vs no screening followed expected patterns.

##### 9-5. Face validity

Model structure, clinical pathways, stage distributions, and treatment patterns were consistent with Japanese lung cancer screening and treatment guidelines [14,23].

**Supplementary Table S1.** EGFR-positive and EGFR-negative odds ratios used for PRS integration

| PRS quartile | EGFR− sample count | EGFR+ sample count | OR (EGFR−) | OR (EGFR+) |
|---|---|---|---|---|
| Quartile 1 | 57 | 33 | 1.00 | 1.00 |
| Quartile 2 | 75 | 80 | 1.29 | 2.52 |
| Quartile 3 | 104 | 115 | 1.81 | 3.85 |
| Quartile 4 | 244 | 290 | 3.50 | 8.63 |

OR values are from multinomial analysis (EGFR− vs controls, EGFR+ vs controls). Sample counts are taken from Blechter et al (2025), Table 1. These values were combined using log-scale weighting (see Supplementary Methods). PRS, polygenic risk scores; EGFR, epidermal growth factor receptor; OR, odds ratio.

**Supplementary Table S2.** Combined PRS–ETS relative risks used in the microsimulation model

| PRS quartile | PRS RR | ETS exposure | ETS RR | Combined RR (PRS × ETS) |
|---|---|---|---|---|
| Quartile 1 | 1.00 | No | 1.00 | 1.00 |

| | | | | |
|---|---|---|---|---|
| Quartile 1 | 1.00 | Yes | 1.33 | 1.33 |
| Quartile 2 | 1.83 | No | 1.00 | 1.83 |
| Quartile 2 | 1.83 | Yes | 1.33 | 2.44 |
| Quartile 3 | 2.68 | No | 1.00 | 2.68 |
| Quartile 3 | 2.68 | Yes | 1.33 | 3.56 |
| Quartile 4 | 5.59 | No | 1.00 | 5.59 |
| Quartile 4 | 5.59 | Yes | 1.33 | 7.43 |

PRS-specific relative risks were derived from log-scale integration of EGFR-positive and EGFR-negative odds ratios (see Supplementary Table S1 and Supplementary Methods). ETS relative risk (RR=1.33) was obtained from Ni et al. (2018). Combined risks assume multiplicative joint effects. These eight strata were used to determine stratum-specific LDCT initiation ages. PRS, polygenic risk scores; ETS, environmental tobacco smoke; RR, relative risk.

**Supplementary Table S3.** Age-specific lung cancer incidence: observed lung cancer vs model lung adenocarcinoma (never-smoking women)

Observed incidence reflects all female lung cancers; modelled incidence reflects lung adenocarcinoma among never-smoking women.

| Age group | Observed female lung cancer incidence (per person-year) | Never-smoking female lung adenocarcinoma incidence (never-smoking women, per person-year) |
|---|---|---|
| 20-24 | 0.000003 | 0.000003 |
| 25-29 | 0.000009 | 0.000008 |
| 30-34 | 0.000017 | 0.000015 |
| 35-39 | 0.000032 | 0.000028 |
| 40-44 | 0.000071 | 0.000062 |
| 45-49 | 0.000128 | 0.000111 |
| 50-54 | 0.000230 | 0.000199 |
| 55-59 | 0.000406 | 0.000350 |
| 60-64 | 0.000668 | 0.000581 |
| 65-69 | 0.001067 | 0.000936 |
| 70-74 | 0.001603 | 0.001418 |
| 75-79 | 0.002010 | 0.001794 |

| | | |
|---|---|---|
| 80-84 | 0.001991 | 0.001777 |
| 85-89 | 0.002007 | 0.001791 |
| 90-94 | 0.002100 | 0.001874 |
| 95-99 | 0.002372 | 0.002117 |
| 100- | 0.002253 | 0.002011 |

**Supplementary Table S4.** Threshold values from one-way sensitivity analyses for strategy switching points in initiation age for annual LDCT in the PRS–ETS model

| Variable | Threshold (RR) | Baseline (years) | Comparator (years) | WTP (US$/QALY) | NMB (US$) |
|---|---|---|---|---|---|
| RR | 7.11 | 40 | 45 | 50,000 | 1,219,952 |
| RR | 2.40 | 45 | 50 | 50,000 | 1,242,018 |
| RR | 1.44 | 50 | 55 | 50,000 | 1,247,005 |

Threshold values indicate the parameter levels at which the optimal LDCT initiation strategy changes. PRS, polygenic risk scores; ETS, environmental tobacco smoke; RR, relative risk; WTP, willingness-to-pay; QALY, quality-adjusted life year; NMB, net monetary benefit.

**Supplementary Table S5.** Optimal screening initiation age by PRS and ETS risk strata

| Risk stratification | Optimal initiation age |
|---|---|
| PRS1 | 55 |
| PRS2 | 50 |
| PRS3 | 45 |
| PRS4 | 45 |
| PRS1+ETS | 55 |
| PRS2+ETS | 45 |
| PRS3+ETS | 45 |
| PRS4+ETS | 40 |

PRS, polygenic risk scores; ETS, environmental tobacco smoke.

**Supplementary Table S6.** Base-case results for the PRS1 stratum

This table provides detailed results supporting Table 2, Figure 3, and Supplementary Figure S1.

| Strategy | Cost (US$) | Incremental Cost (US$) | Effectiveness (QALY) | Incremental Effectiveness (QALY) | ICER (US$/QALY) | NMB (US$) |
|---|---|---|---|---|---|---|
| No screening | 3,298 | - | 20.6037 | - | - | 1,026,889 |
| LDCT every 10 years | 3,617 | 319 | 20.6080 | 0.0042 | 75,460 | 1,026,781 |
| LDCT every 5 years | 3,684 | 67 | 20.6117 | 0.0038 | 17,807 | 1,026,903 |
| LDCT every 4 years | 3,718 | 33 | 20.6137 | 0.0019 | 17,231 | 1,026,966 |
| LDCT every 3 years | 3,775 | 57 | 20.6168 | 0.0031 | 18,111 | 1,027,066 |
| Annual CXR | 3,342 | -432 | 20.6202 | 0.0033 | dominant | 1,027,666 |
| LDCT every 2 years | 3,888 | 546 | 20.6231 | 0.0030 | 183,862 | 1,027,269 |
| Annual LDCT | 4,228 | 340 | 20.6421 | 0.0189 | 17,985 | 1,027,875 |

“Dominant” indicates an intervention that is both more effective and less costly than the comparator. PRS, polygenic risk scores; LDCT, low-dose computed tomography; CXR, chest radiography; ICER, incremental cost-effectiveness ratio; NMB, net monetary benefit.

**Supplementary Table S7.** Base-case results for the PRS2 stratum

This table provides detailed results supporting Table 2, Figure 3, and Supplementary Figure S1.

| Strategy | Cost (US$) | Incremental Cost (US$) | Effectiveness (QALY) | Incremental Effectiveness (QALY) | ICER (US$/QALY) | NMB (US$) |
|---|---|---|---|---|---|---|
| No screening | 5,418 | - | 22.1418 | - | - | 1,101,672 |

| | | | | | | |
|---|---|---|---|---|---|---|
| LDCT every 10 years | 5,707 | 289 | 22.1487 | 0.0070 | 41,569 | 1,101,730 |
| LDCT every 5 years | 5,743 | 36 | 22.1553 | 0.0066 | 5,423 | 1,102,023 |
| LDCT every 4 years | 5,760 | 18 | 22.1586 | 0.0033 | 5,293 | 1,102,172 |
| LDCT every 3 years | 5,791 | 31 | 22.1641 | 0.0054 | 5,774 | 1,102,411 |
| Annual CXR | 5,312 | -480 | 22.1700 | 0.0060 | dominant | 1,103,189 |
| LDCT every 2 years | 5,853 | 541 | 22.1750 | 0.0049 | 109,472 | 1,102,895 |
| Annual LDCT | 6,038 | 185 | 22.2076 | 0.0326 | 5,679 | 1,104,342 |

“Dominant” indicates an intervention that is both more effective and less costly than the comparator. PRS, polygenic risk scores; LDCT, low-dose computed tomography; CXR, chest radiography; ICER, incremental cost-effectiveness ratio; NMB, net monetary benefit.

**Supplementary Table S8.** Base-case results for the PRS3 stratum

This table provides detailed results supporting Table 2, Figure 3, and Supplementary Figure S1.

| **Strategy** | **Cost (US$)** | **Incremental Cost (US$)** | **Effectiveness (QALY)** | **Incremental Effectiveness (QALY)** | **ICER (US$/QALY)** | **NMB (US$)** |
|---|---|---|---|---|---|---|
| No screening | 6,963 | - | 23.5351 | - | - | 1,169,791 |
| LDCT every 10 years | 7,231 | 268 | 23.5442 | 0.0091 | 29,454 | 1,169,978 |
| LDCT every 5 years | 7,247 | 16 | 23.5528 | 0.0086 | 1,883 | 1,170,393 |
| LDCT every 4 years | 7,255 | 8 | 23.5572 | 0.0044 | 1,745 | 1,170,606 |
| LDCT every 3 years | 7,267 | 12 | 23.5645 | 0.0073 | 1,699 | 1,170,959 |
| Annual CXR | 6,751 | -516 | 23.5725 | 0.0080 | dominant | 1,171,876 |

| | | | | | | |
|---|---|---|---|---|---|---|
| LDCT every 2 years | 7,294 | 543 | 23.5790 | 0.0065 | 84,067 | 1,171,656 |
| Annual LDCT | 7,375 | 81 | 23.6224 | 0.0434 | 1,864 | 1,173,746 |

"Dominant" indicates an intervention that is both more effective and less costly than the comparator. PRS, polygenic risk scores; LDCT, low-dose computed tomography; CXR, chest radiography; ICER, incremental cost-effectiveness ratio; NMB, net monetary benefit.

**Supplementary Table S9.** Base-case results for the PRS4 stratum

This table provides detailed results supporting Table 2, Figure 3, and Supplementary Figure S1.

| **Strategy** | **Cost (US$)** | **Incremental Cost (US$)** | **Effectiveness (QALY)** | **Incremental Effectiveness (QALY)** | **ICER (US$/QALY)** | **NMB (US$)** |
|---|---|---|---|---|---|---|
| No screening | 13,807 | - | 23.2481 | - | - | 1,148,596 |
| LDCT every 10 years | 13,937 | 131 | 23.2664 | 0.0183 | 7,139 | 1,149,381 |
| LDCT every 5 years | 13,822 | -116 | 23.2837 | 0.0174 | dominant | 1,150,365 |
| LDCT every 4 years | 13,762 | -59 | 23.2926 | 0.0089 | dominant | 1,150,869 |
| LDCT every 3 years | 13,663 | -99 | 23.3073 | 0.0147 | dominant | 1,151,702 |
| Annual CXR | 13,033 | -631 | 23.3234 | 0.0161 | dominant | 1,153,138 |
| LDCT every 2 years | 13,469 | 436 | 23.3364 | 0.0130 | 33,496 | 1,153,353 |
| Annual LDCT | 12,885 | -583 | 23.4238 | 0.0874 | dominant | 1,158,305 |

"Dominant" indicates an intervention that is both more effective and less costly than the comparator. PRS, polygenic risk scores; LDCT, low-dose computed tomography; CXR, chest radiography; ICER, incremental cost-effectiveness ratio; NMB, net monetary benefit.

**Supplementary Table S10.** Base-case results for the PRS1+ETS stratum

This table provides detailed results supporting Table 2, Figure 3, and Supplementary Figure S1.

| Strategy | Cost (US$) | Incremental Cost (US$) | Effectiveness (QALY) | Incremental Effectiveness (QALY) | ICER (US$/QALY) | NMB (US$) |
|---|---|---|---|---|---|---|
| No screening | 4,368 | - | 20.5630 | - | - | 1,023,781 |
| LDCT every 10 years | 4,665 | 297 | 20.5686 | 0.0056 | 52,906 | 1,023,765 |
| LDCT every 5 years | 4,712 | 47 | 20.5736 | 0.0050 | 9,313 | 1,023,969 |
| LDCT every 4 years | 4,735 | 23 | 20.5762 | 0.0026 | 8,891 | 1,024,074 |
| LDCT every 3 years | 4,774 | 40 | 20.5803 | 0.0042 | 9,544 | 1,024,243 |
| Annual CXR | 4,325 | -450 | 20.5848 | 0.0044 | dominant | 1,024,915 |
| LDCT every 2 years | 4,853 | 529 | 20.5887 | 0.0039 | 134,215 | 1,024,583 |
| Annual LDCT | 5,090 | 237 | 20.6138 | 0.0251 | 9,442 | 1,025,601 |

“Dominant” indicates an intervention that is both more effective and less costly than the comparator. PRS, polygenic risk scores; ETS, environmental tobacco smoke; LDCT, low-dose computed tomography; CXR, chest radiography; ICER, incremental cost-effectiveness ratio; NMB, net monetary benefit.

**Supplementary Table S11.** Base-case results for the PRS2+ETS stratum

This table provides detailed results supporting Table 2, Figure 3, and Supplementary Figure S1.

| Strategy | Cost (US$) | Incremental Cost (US$) | Effectiveness (QALY) | Incremental Effectiveness (QALY) | ICER (US$/QALY) | NMB (US$) |
|---|---|---|---|---|---|---|
| Annual CXR | 6,199 | - | 23.5941 | - | - | 1,173,505 |
| No screening | 6,361 | 163 | 23.5599 | -0.0342 | -4,761 | 1,171,634 |

| | | | | | | |
|---|---|---|---|---|---|---|
| LDCT every 10 years | 6,641 | 442 | 23.5682 | -0.0259 | -17,099 | 1,171,769 |
| LDCT every 5 years | 6,669 | 470 | 23.5761 | -0.0180 | -26,130 | 1,172,135 |
| LDCT every 4 years | 6,683 | 484 | 23.5801 | -0.0140 | -34,638 | 1,172,323 |
| LDCT every 3 years | 6,705 | 506 | 23.5868 | -0.0073 | -69,197 | 1,172,633 |
| LDCT every 2 years | 6,751 | 552 | 23.6000 | 0.0059 | 93,733 | 1,173,247 |
| Annual LDCT | 6,891 | 692 | 23.6396 | 0.0455 | 15,197 | 1,175,090 |

"Dominant" indicates an intervention that is both more effective and less costly than the comparator. PRS, polygenic risk scores; ETS, environmental tobacco smoke; LDCT, low-dose computed tomography; CXR, chest radiography; ICER, incremental cost-effectiveness ratio; NMB, net monetary benefit.

**Supplementary Table S12.** Base-case results for the PRS3+ETS stratum

This table provides detailed results supporting Table 2, Figure 3, and Supplementary Figure S1.

| Strategy | Cost (US$) | Incremental Cost (US$) | Effectiveness (QALY) | Incremental Effectiveness (QALY) | ICER (US$/QALY) | NMB (US$) |
|---|---|---|---|---|---|---|
| No screening | 9,130 | - | 23.4451 | - | - | 1,163,126 |
| LDCT every 10 years | 9,354 | 224 | 23.4571 | 0.0120 | 18,734 | 1,163,501 |
| LDCT every 5 years | 9,329 | -26 | 23.4685 | 0.0114 | dominant | 1,164,095 |
| LDCT every 4 years | 9,315 | -13 | 23.4743 | 0.0058 | dominant | 1,164,399 |
| LDCT every 3 years | 9,292 | -23 | 23.4839 | 0.0096 | dominant | 1,164,903 |
| Annual CXR | 8,740 | -552 | 23.4944 | 0.0106 | dominant | 1,165,983 |

| | | | | | | |
|---|---|---|---|---|---|---|
| LDCT every 2 years | 9,249 | 509 | 23.5030 | 0.0085 | 59,821 | 1,165,899 |
| Annual LDCT | 9,120 | -129 | 23.5602 | 0.0572 | dominant | 1,168,889 |

"Dominant" indicates an intervention that is both more effective and less costly than the comparator. PRS, polygenic risk scores; ETS, environmental tobacco smoke; LDCT, low-dose computed tomography; CXR, chest radiography; ICER, incremental cost-effectiveness ratio; NMB, net monetary benefit.

**Supplementary Table S13.** Base-case results for the PRS4+ETS stratum

This table provides detailed results supporting Table 2, Figure 3, and Supplementary Figure S1.

| Strategy | Cost (US$) | Incremental Cost (US$) | Effectiveness (QALY) | Incremental Effectiveness (QALY) | ICER (US$/QALY) | NMB (US$) |
|---|---|---|---|---|---|---|
| No screening | 15,682 | - | 24.4487 | - | - | 1,206,754 |
| LDCT every 10 years | 15,790 | 107 | 24.4697 | 0.0209 | 5,126 | 1,207,693 |
| LDCT every 5 years | 15,643 | -147 | 24.4901 | 0.0204 | dominant | 1,208,862 |
| LDCT every 4 years | 15,568 | -74 | 24.5006 | 0.0105 | dominant | 1,209,460 |
| LDCT every 3 years | 15,446 | -122 | 24.5177 | 0.0172 | dominant | 1,210,440 |
| Annual CXR | 14,773 | -673 | 24.5368 | 0.0191 | dominant | 1,212,067 |
| LDCT every 2 years | 15,204 | 431 | 24.5518 | 0.0150 | 28,723 | 1,212,387 |
| Annual LDCT | 14,476 | -727 | 24.6542 | 0.1024 | dominant | 1,218,232 |

"Dominant" indicates an intervention that is both more effective and less costly than the comparator. PRS, polygenic risk scores; ETS, environmental tobacco smoke; LDCT, low-dose computed tomography; CXR, chest radiography; ICER, incremental cost-effectiveness ratio; NMB, net monetary benefit.

**Supplementary Table S14.** Threshold Values Identified in One-Way Sensitivity Analysis for the PRS1 Stratum

| Parameter | Low | Base | High | Threshold Value | Impact | Interpretation |
|---|---|---|---|---|---|---|
| Discount rate (annual) | 0 | 0.03 | 0.05 | 0.043 | Increase | NMB negative above 0.043 |
| Stage I proportion under LDCT screening | 0.8 | 0.961 | 0.97 | 0.911 | Decrease | NMB worsens below 0.911 |
| LDCT unit cost | 79 | 106 | 132 | 122.2 | Increase | NMB worsens above 122.2 |
| Stage I proportion without screening | 0.45 | 0.539 | 0.6 | 0.589 | Increase | Benefit declines above 0.589 |
| Adherence: LDCT → CXR pathway | 0.5 | 0.7 | 0.9 | 0.834 | Increase | NMB improves above 0.834 |
| LDCT adherence rate | 0.5 | 0.6 | 0.8 | — | Decrease | — |
| CXR adherence rate | 0.4 | 0.5 | 0.6 | — | Increase | — |
| Stage IV treatment cost (per case) | 79,365 | 105,820 | 132,275 | — | Decrease | — |

PRS, polygenic risk scores; LDCT, low-dose computed tomography; CXR, chest radiography; NMB, net monetary benefit.

**Supplementary Table S15.** Optimal LDCT initiation age under ETS relative risk scenarios (−20%, baseline, +20%)

| Risk stratum | Base ETS (years) | Household ETS (years) | Workplace ETS (years) | Combined ETS (years) |
|---|---|---|---|---|
| PRS1+ETS | 55 / 55 / 50 | 55 / 50 / 50 | 55 / 50 / 50 | 50 / 45 / 45 |
| PRS2+ETS | 50 / 45 / 45 | 50 / 45 / 45 | 45 / 45 / 45 | 45 / 45 / 45 |
| PRS3+ETS | 45 / 45 / 45 | 45 / 45 / 45 | 45 / 45 / 45 | 45 / 40 / 40 |
| PRS4+ETS | 45 / 40/ 40 | 45 / 40 / 40 | 40/ 40/ 40 | 40/ 40 / 40 |

Base ETS reflects three scenarios (−20%, baseline, +20%). Household ETS, Workplace ETS, and Combined ETS represent alternative exposure-specific relative risk assumptions. LDCT, low-dose computed tomography; ETS, environmental tobacco smoke; PRS, polygenic risk scores.

**Supplementary Table S16.** Population-level lifetime costs and health outcomes for annual LDCT versus annual CXR and versus no screening

| **Outcome** | **Annual LDCT vs annual CXR** | **Annual LDCT vs No screening** |
|---|---|---|
| Incremental cost, US$ | 158,964,599 | 58,518,931 |
| Incremental QALYs | 51,616 | 64,991 |
| Stage I cancers detected (additional), n | 50,777 | 88,888 |
| Stage II cancers prevented, n | 6,385 | 11,177 |
| Stage III cancers prevented, n | 6,918 | 12,110 |
| Stage IV cancers prevented, n | 17,114 | 29,959 |
| LAC-related deaths averted, n | 8,534 | 14,940 |

Population estimates vary by risk stratum and optimal LDCT initiation age, as determined in Table S2. LDCT, low-dose computed tomography; CXR, chest radiography; QALY, quality-adjusted life year; LAC, lung adenocarcinoma.

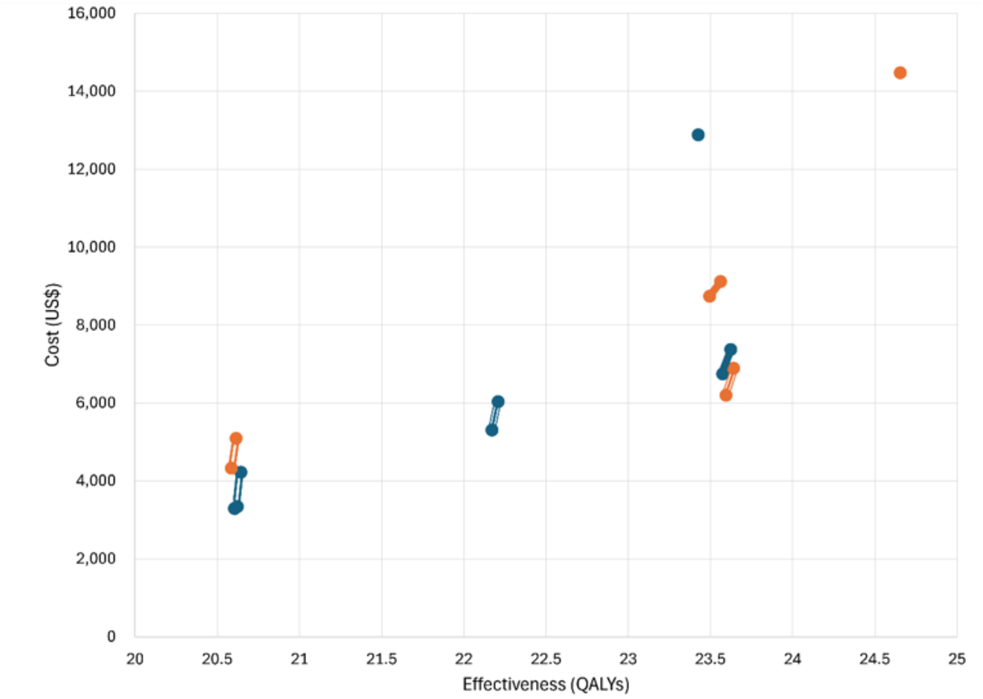


**Supplementary Figure S1.** Efficiency frontier showing the cost–effectiveness positions of annual LDCT and annual CXR across combinations of polygenic risk score (PRS) levels and environmental tobacco smoke (ETS) exposure.

Blue and orange markers indicate ETS-absent and ETS-present groups, respectively. Line styles represent PRS levels: thin solid line (2-stroke) for PRS1, medium solid line (3-stroke) for PRS2, thick solid line for PRS3, and markers only for PRS4. LDCT, low-dose computed tomography; CXR, chest radiography.

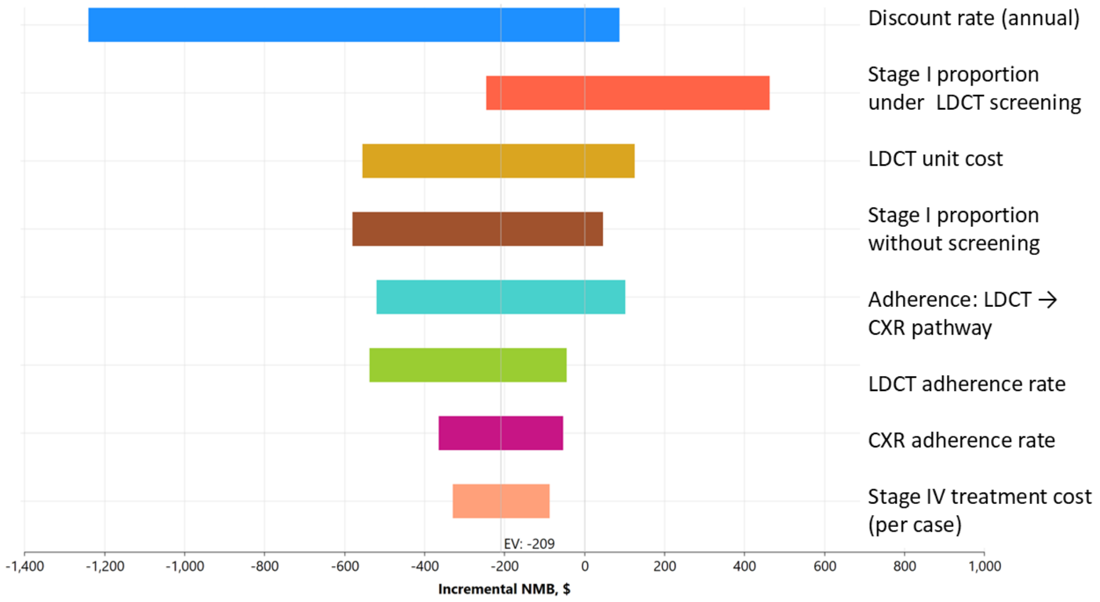


**Supplementary Figure S2.** One-way sensitivity analysis showing the parameters with the greatest impact on the incremental cost-effectiveness ratio (ICER) of annual LDCT screening in the PRS1 stratum.

Parameters with minimal impact on incremental NMB were excluded from the tornado diagram for clarity. LDCT, low-dose computed tomography; CXR, chest radiography.
